# Diet-derived antioxidants do not decrease the risk of ischemic stroke: a Mendelian Randomization Study in over 1 million participants

**DOI:** 10.1101/2021.03.19.21253362

**Authors:** Leon G Martens, Jiao Luo, Ko Willems van Dijk, J Wouter Jukema, Raymond Noordam, Diana van Heemst

**Affiliations:** Department of Internal Medicine, Section of Gerontology and Geriatrics, Leiden University Medical Center, Leiden, the Netherlands; Department of Clinical Epidemiology, Leiden University Medical Center, Leiden, the Netherlands; Department of Human Genetics, Leiden University Medical Center, Leiden, the Netherlands; Department of Internal Medicine, Division of Endocrinology, Leiden University Medical Center, Leiden, the Netherlands; Einthoven Laboratory for Experimental Vascular Medicine, Leiden University Medical Center, Leiden, the Netherlands; Department of Cardiology, Leiden University Medical Center, Leiden, the Netherlands; Netherlands Heart Institute, Utrecht, the Netherlands

**Author notes:** Address of correspondence Leon G Martens MSc, Department of Internal Medicine, Section of Gerontology and Geriatrics, Leiden University Medical Center, PO Box 9600, 2300 RC, Leiden, the Netherlands. Contributed equally (Leon G Martens, Jiao Luo).

## Abstract

**Background:** Intake, either as dietary components or as supplements, and blood concentrations of vitamin E, C, lycopene and carotenoids have been associated with a lower risk of incident (ischemic) stroke. However, causality cannot not be inferred from these associations. Here, we investigated causality by analyzing the associations between genetically-influenced antioxidant levels in blood and ischemic stroke using Mendelian Randomization (MR).

**Methods:** For each circulating antioxidant (vitamin E, C, lycopene, β-carotene and retinol), which were assessed as either absolute blood levels and/or high-throughput metabolite levels, genetic instrumental variables were generated from earlier genome wide association studies. We used summary statistics for SNP-stroke associations from three European-ancestry cohorts (cases/controls): MEGASTROKE (67,162/454,450), UK Biobank (2,404/368,771) and FinnGen study (8,046/164,286). MR analyses were performed on each exposure per outcome cohort using inverse-variance weighted analyses, and subsequently meta-analyzed.

**Results:** In a combined sample of 1,065,119 individuals (77,612 cases), none of the genetically-influenced absolute antioxidants or antioxidant metabolite concentrations were causally associated with a lower risk of ischemic stroke. For absolute antioxidants levels, the odds ratios (95% CI) ranged between 0.94 (95% CI: 0.85 to 1.05) for vitamin C and 1.04 (95% CI: 0.99 to 1.08) for lycopene. For metabolites, odds ratios ranged between 1.01 (95% CI: 0.98 to 1.03) for retinol and 1.12 (95% CI: 0.88 to 1.42) for vitamin E.

**Conclusion:** This study did not provide evidence for a causal association between dietary-derived antioxidant levels and ischemic stroke. Therefore, antioxidant supplements to increase circulating levels are unlikely to be of clinical benefit to prevent ischemic stroke.

## Introduction

Stroke is the second leading cause of death and loss of disability-adjusted life years worldwide.[1] Stroke is characterized by a sudden deficit of neurological origin, and is most frequently (about 80%) caused by a disruption of cerebral blood flow causing a lack of oxygen in the affected area, and is therefore referred to as ischemic stroke.[2] Several risk factors have been described as important in the pathogenesis of ischemic stroke, including smoking, obesity, diabetes mellitus, hypertension and dyslipidemia [3-7]. However, interventions aiming to reduce the global burden of stroke are challenging. In addition to the traditional risk factors, oxidative stress has been hypothesized to be a vital trigger in the occurrence of stroke via an excess production of Reactive Oxygen Species (ROS).[8, 9] ROS-induced damage can cause significant changes in the vascular system, ultimately influencing cerebral blood flow.[9] These detrimental effects, including increasing vasodilation, platelet aggregation, increased endothelial permeability and the formation of local lesions, [9] could consequently lead to an increased risk of stroke.[10]

Therefore, antioxidants, which are scavengers of free radicals and thereby diminish oxidative damage, can be hypothesized to decrease the risk of disease occurrence. Antioxidants, such as vitamin E, C and carotenoids, are of specific interest given that they are accessible and their intake is easily modifiable. Several studies have already been conducted examining the association between antioxidants and the occurrence of stroke.[8, 10-14] Here, dietary intake, either as dietary components or supplements, or blood concentration of vitamin E, C and carotenoids were associated with lower risk of first occurrence of ischemic stroke.[8, 11-14] Similarly, adherence to a diet rich in antioxidants, irrespective of the type of antioxidants, has been associated with a lower risk of incident stroke.[15]

However, associations in these observational studies inevitably suffer from the possibility of reverse causation and residual confounding, and should be interpreted with caution. Therefore, the causality between dietary-derived antioxidants and stroke is still unclear. A randomized controlled trial on vitamin E and β-carotene supplementation and stroke risk found inconsistent results.[16] Whereas a similar study on vitamin E supplementation and cardiovascular events found no effect.[17] In addition to randomized clinical trials, Mendelian Randomization (MR), in which genetic variants of a certain exposure are used as instrumental variables, is an alternative approach to infer causality of life-long risk factors (exposure) on diseases (outcome).[18-20] In the present study, we used two-sample MR to assess the associations between genetically-influenced dietary-derived circulating antioxidants and their metabolites with ischemic stroke.

## Methods

### Selection of genetic instrumental variables

To study the association between genetically-influenced levels of antioxidants and stroke, the genetic instruments of five diet-derived antioxidants were used, which included vitamin E (α-tocopherol and γ-tocopherol), β-carotene, lycopene, vitamin C (L-ascorbic acid or ascorbate) and retinol. For these antioxidants, we investigated either absolute blood levels, circulating metabolites which were quantified as relative concentrations in plasma or serum using high-throughput commercial platforms, or both. The metabolites were quantified as relative concentrations.[21] For each antioxidant, a search for Genome Wide Association Studies (GWAS) was performed to extract the leading SNPs as genetic instrumental variables. Whenever multiple GWAS were identified, only the largest study including replication was used.[22-28] A summary table of the SNPs for each antioxidant used as instrumental variables can be found in our previous publication.[21] Different from our earlier work[21], we now used an updated genome-wide association study on vitamin C (genetic instruments presented in **Supplementary Table 1**).[28]

**Table 1:** GWAS on genetically determined dietary derived antioxidants

| <u>Exposure Datasets</u> | <u>N =</u> | <u>Absolute antioxidants</u> |  | <u>Unit</u> | <u>N=</u> | <u>Metabolite antioxidants</u> |  | <u>Units</u> |
| --- | --- | --- | --- | --- | --- | --- | --- | --- |
|  |  | <u>Genetic Variants</u> | <u>Explained Variance</u> |  |  | <u>Genetic Variants</u> | <u>Explained Variance</u> |  |
| a-Tocopherol | - | - | - | - | 7276 | 11 | 3.3 | log10-transformed metabolites concentration |
| γ-Tocopherol | - | - | - | - | 5822 | 13 | 15.0% | log10-transformed metabolites concentration |
| β-Carotene | 2344 | 3 | 9.0% | μg/L in log-transformed scale | - | - | - | - |
| Lycopene | 441 | 5 | 30.1% | μg/dL | - | - | - | - |
| Ascorbate | 52018 | 11 | 1.87% | μmol/L | 2063 | 13 | 18.6% | log10-transformed metabolites concentration |
| Retinol | 5006 | 2 | 2.3% | μg/L in log-transformed scale | 1957 | 24 | 4.8% | log10-transformed metabolites concentration |

Variance explained (R^2^) by the instruments for each trait were either derived from the original study or calculated based on the derived summary statistics and in line with the method described previously [29], and ranged from 1.7% to 30.1% for absolute antioxidant levels, and from 3.3% to 18.6% for metabolite antioxidants (**Table 1**). In order to minimize potential weak instrument bias, we considered an F-statistic of at least 10 as sufficient for performing an MR analysis, which is well-accepted in the field.[30]

### Outcome datasets

Summary statistics on the associations of the exposure-related SNPs with ischemic stroke were extracted from three large cohorts; the MEGASTROKE consortium, the UK Biobank, and the FinnGen study. Both UK Biobank and FinnGen were not part of the main analyses of the MEGASTROKE consortium preventing inclusion of overlapping samples in the analyses.

The MEGASTROKE consortium consisted of 67,162 cases and 454,450 controls collected from 29 studies. Of these participants, 86% were of European ancestry, 9% of East Asian ancestry, and the remaining participants were from African, South Asian, mixed Asian, and Latin American ancestry.[31]

The UK Biobank cohort (project application number 56340) is a prospective general population cohort with 502,628 participants between the age of 40 and 70 years recruited from the general population between 2006 and 2010 [32], and more information can be found online (https://www.ukbiobank.ac.uk). The analyses were performed with participants of European ancestry, who were in the full released imputed genomics databases (UK10K + HRC). Follow-up information, including stroke occurrence, was retrieved through routinely available national datasets. In our dataset, we had data available on 2,404 cases of ischemic stroke, and 368,771 controls. We performed new genome-wide association analyses using logistic regression to assess the associations between genetic instruments and ischemic stroke, adjusted for age, sex and 10 principal components, and corrected for familial relationships using BOLT_LMM (v2.3.2).

The FinnGen study is an ongoing cohort study launched in 2017, which integrated the genetic data generated from biobank samples and health-related data from social and healthcare registers. Detailed information such as participating biobanks/cohorts, genotyping and data analysis are available at their website (https://www.finngen.fi/en/). For our current study, the freeze 4 data was used. Within this data, there are 8,046 reported cases of ischemic stroke, and 164,286 controls.

### Statistical analysis

All the analyses were done using R (v3.6.1) statistical software (The R Foundation for Statistical Computing, Vienna, Austria). MR analyses were performed using the R-based package “TwoSampleMR” (https://mrcieu.github.io/TwoSampleMR/).

For our primary MR analysis, Inverse-Variance weighted (IVW) regression analyses were performed. This method assumes the absence of invalid genetic instruments such as SNPs affecting multiple exposures (pleiotropy) causing possible directional pleiotropy.[19] First, causal estimates were calculated per genetic instrument using the Wald ratio (SNP – outcome association divided by the SNP – exposure association) and subsequently meta-analyzed using the inverse-weighted meta-analyses weighted on the standard error of the SNP-outcome association (assuming no measurement error [NOME] in the exposure).[33] The calculated estimates were expressed as odds ratios (OR) on ischemic stroke per unit difference of the corresponding absolute circulating antioxidant levels (natural log-transformed levels for β-carotene and retinol, µg/dL for lycopene or μmol/L for ascorbate) or 10-fold change in antioxidant metabolite concentrations.

To ensure that the results obtained from the IVW analyses were not biased due to directional pleiotropy, we performed MR-Egger regression analysis and Weighted-Median Estimator when applicable.[33] In MR-Egger, the intercept depicts the estimated average pleiotropic effect across the genetic variants, and a value that differs from zero indicates that the IVW estimate is biased [34]. Although considered as a relatively inefficient approach (e.g., large confidence intervals), this method does not force the regression line to go through the intercept. The Weighted-Median estimator analysis can provide a consistent valid estimate if at least half of the instrumental variables are valid [20]. In addition, MR-PRESSO (MR Pleiotropy RESidual Sum and Outlier) was applied when possible to detect and correct for horizontal pleiotropy through removing outliers [35], as implemented in the R-based package MRPRESSO (https://github.com/rondolab/MR-PRESSO)*”*. The Cochran’s Q statistic was performed in order to test the heterogeneity between the estimated Wald ratios from different genetic variants. [36] Additional sensitivity analyses were performed for the antioxidants β-carotene and lycopene.

The main MR analyses were performed in the individual datasets, and subsequently meta-analyzed to derive the pooled estimates for each exposure on the risk of ischemic stroke. Heterogeneity of the estimates across three datasets was performed by I^2^, and corresponding p-value was obtained from the Cochran’s Q test. When no heterogeneity was found amongst the three cohorts, a fixed-effect model meta-analysis was used to pool the instrumental variable estimates for each exposure. All meta-analyses were performed in the R-based “meta” package (https://cran.r-project.org/web/packages/meta/index.html).

## Results

By combining the three cohorts, a total sample of 1,065,119 individuals, of which 77,612 cases of ischemic stroke, and 987,507 controls, were analyzed to assess the association between diet-derived antioxidants and ischemic stroke. The in-between SNP heterogeneity was non-significant for all antioxidants in each cohort (p > 0.05). Additionally, we found no heterogeneity in the summary estimates from the MR analyses between the included datasets (**Figure 1 and Figure 2**).

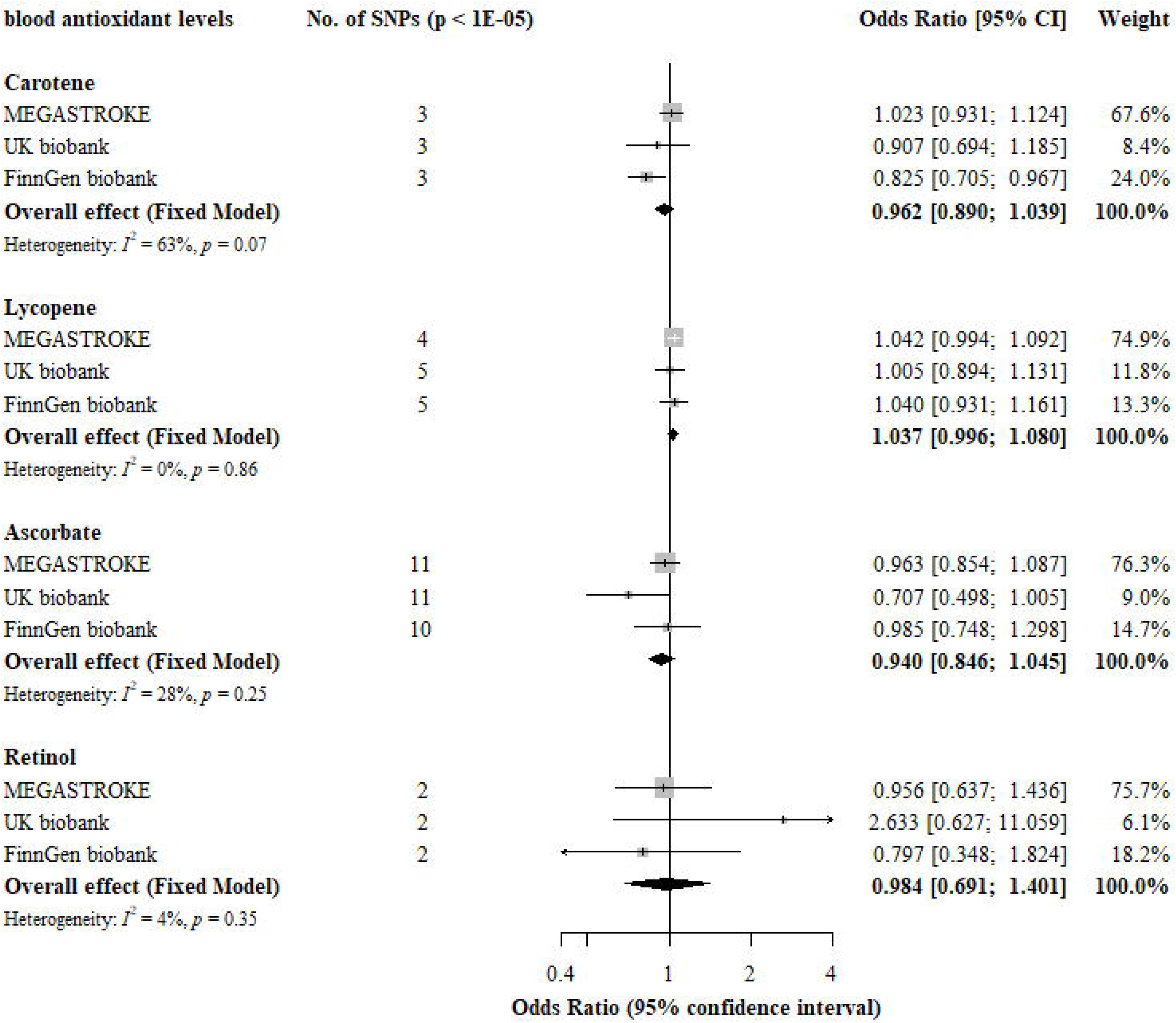

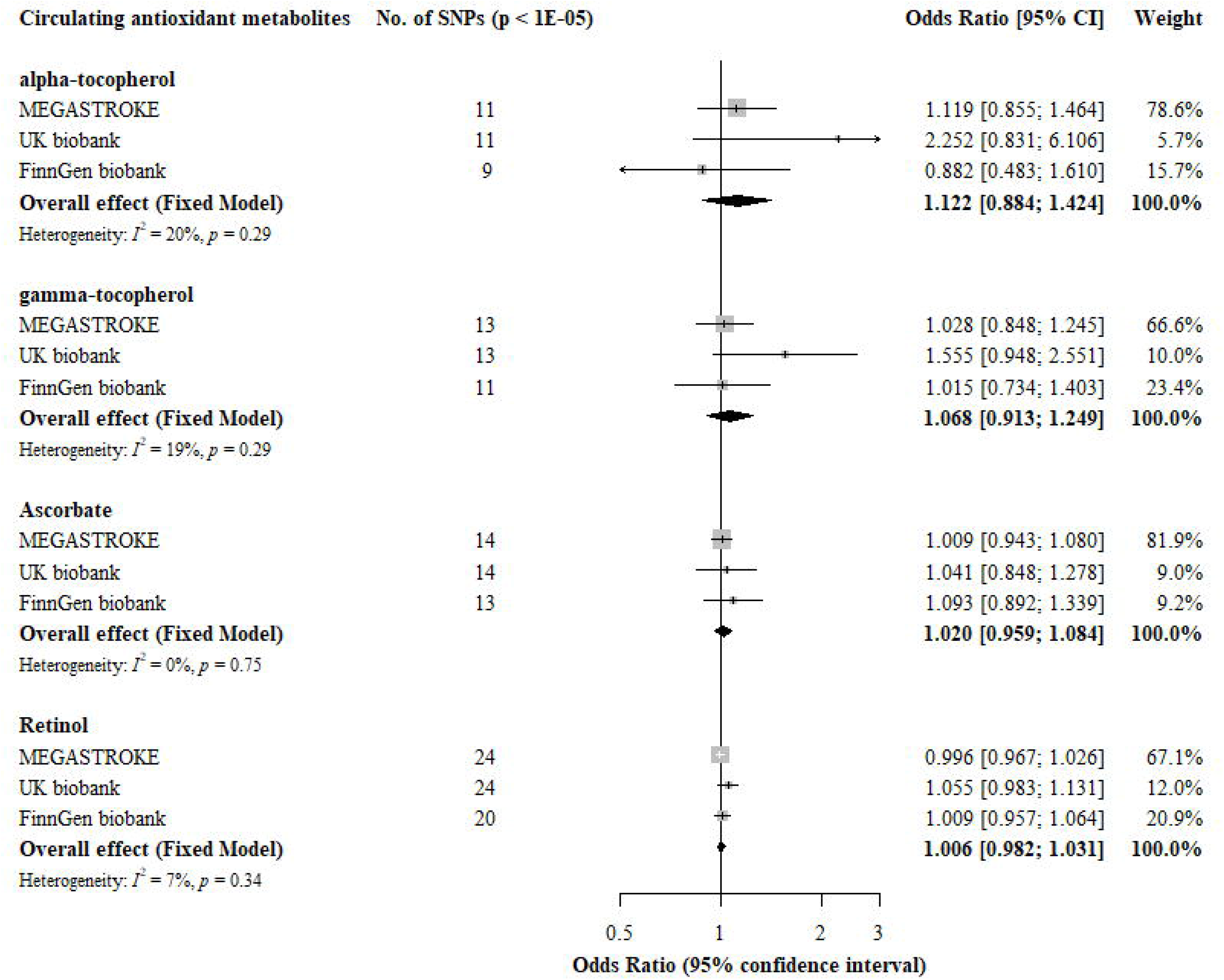

### Absolute antioxidant levels

For absolute blood antioxidants levels (**Figure 1**), we observed no evidence for associations with ischemic stroke, except for β-carotene when analyzed only within the FinnGen cohort. However, the pooled estimate for all three cohorts was not significantly associated with ischemic stroke. Most notably, the pooled odds ratios (95% confidence intervals) were 0.94 (95% CI: 0.85, 1.05) per 1 µmol/L ascorbate, 1.04 (95% CI: 0.99, 1.08) per 1 µg/dL lycopene, 0.96 (95% CI: 0.89, 1.04) and 0.98 (95% CI: 0.69, 1.40) per natural log-transformed β-carotene and retinol, respectively.

MR-Egger and weighted-median estimator regression were performed for antioxidants with more than three genetic instruments (notably β-carotene and lycopene). The estimates of both MR-Egger and weighted-median estimator were comparable with the IVW analyses. Furthermore, the MR-Egger intercept did not deviate from zero (p-values>0.05). Additionally, MR-PRESSO did not detect any outliers, and Cochran’s Q statistics detected no heterogeneity for the analyses of β-carotene or lycopene on ischemic stroke.

### Circulating antioxidant metabolite levels

For circulating antioxidant metabolites (**Figure 2**), the pooled OR (95%CI) for ischemic stroke per 10-fold increase in metabolite concentration were 1.12 (95% CI: 0.88, 1.42) for α - tocopherol, 1.07 (95% CI: 0.91, 1.25) for γ-tocopherol, 1.02 (95% CI: 0.96, 1.08) for ascorbate and 1.01 (95% CI: 0.98, 1.03) for retinol using IVW.

Estimates derived from the MR-Egger and weighted-median estimator analyses were of similar direction and magnitude as the IVW analyses. Furthermore, no pleiotropic effect was identified by the intercept from MR-Egger or MR PRESSO, and no potential outlier was found via MR PRESSO. Cochran’s Q statistics only detected heterogeneity in γ-tocopherol in the MEGASTROKE cohort.

## Discussion

We investigated whether we could find evidence supporting causality of the association between diet-derived antioxidants and ischemic stroke risk using MR. Circulating antioxidants, irrespective of how the levels were determined, were proxied by using genetic variants as instrumental variables. In an extreme sample size of 1,065,119 participants, 77,612 cases and 987,507 controls, we did not find evidence that genetically-influenced diet-derived antioxidant levels were associated with a lower risk of developing ischemic stroke. These findings suggest that the previously observed association between antioxidants (either by dietary intake and/or serum levels) and ischemic stroke is not causal.

Previously, a meta-analysis of observational studies identified a 17% risk reduction of stroke in participants with high dietary vitamin E intake compared with those with low intake. However, the authors were cautious with interpreting this result given that only 3,284 events of ischemic stroke were reported, despite the study comprising over 220,000 participants, as well as the large heterogeneity of the individual contributing studies in the meta-analysis.[37] To date, there are no MR studies that have assessed the causal association of circulating antioxidants and ischemic stroke risk. In our previous study, we demonstrated that the effect of genetic variants on circulating antioxidant levels are generally comparable with those which would be achieved by dietary supplementation [21]. Given the robust and generally consistent null results in the present study that both absolute blood antioxidant levels and metabolites measured with high throughput technology are not causally associated with ischemic stroke, it is not likely that dietary supplements that are to increase antioxidant concentrations in blood reduce the risk of ischemic stroke.

Despite the use of data from more than 1 million participants, our study population could be seen as relatively young for stroke occurrence (especially in the UK Biobank population who were below age 70 years at study inclusion). However, importantly, our findings were robust across different analysis methods and were generally consistent across three independent cohorts with a large number of cases. As our null findings were generally consistent across our study cohorts, it is very unlikely that changes in antioxidant levels will yield any clinically relevant reduction in ischemic stroke risk. These findings were also in line with our earlier work in which we found no associations between these antioxidants in relation to coronary heart disease.[21] Together with our present findings, this would suggest that antioxidants do not affect the risk of developing atherogenic cardiovascular disease. However, this is contrary to the data from experimental settings in which oxidative stress does play an important role in the onset of atherosclerosis.[38, 39] This has given rise to the hypothesis that circulating antioxidant levels might not be representative of antioxidant capacity, and that increasing antioxidant levels in blood (either by nutritional intake or supplements) do not necessarily result in additional antioxidative effects. This hypothesis was supported by a number of our earlier studies in which we investigated the associations of vitamin E and its enzymatic and oxidative metabolites with lifestyle factors and subclinical disease outcomes [40, 41]. In brief, we showed in these studies that vitamin E concentrations were not correlated with the urinary enzymatic and oxidative vitamin E metabolite levels, and lifestyle factors and subclinical disease outcomes showed different associations with vitamin E concentrations than with their oxidized metabolites.

The present study has several strengths. First, a large sample size was studied, by combining three cohorts comprising a total of 77,612 cases and performing a meta-analysis. Individually, the results from the three cohorts are consistent with each other and with the final meta-analysis. We did not detect any heterogeneity between-SNPs for every antioxidant in each cohort, or across each cohort. Additionally, by performing MR-egger and weighted-median estimator analyses, the final MR estimates should be seen as a reliable result despite the sometimes low explained variance of certain genetic instruments. Second, we used separate sets of instrumental variables by looking at both absolute blood levels as well as metabolite levels of antioxidants. The similar findings, especially in regards to ascorbate and retinol which are analyzed with both their absolute blood concentration and the relative metabolite levels, suggests general robustness of our findings. However, some limitations should also be considered with respect to the interpretation of the results. First, participants included in our study are predominantly of European descent, which will limit the extrapolation to other populations. Second, sensitivity analyses for some instrumental variables with limited number of genetic variants as instrumental variables could not be performed.

In summary, our study did not provide evidence supporting a causal association between diet-derived levels of antioxidants vitamin E, C, lycopene, β-carotene and retinol, and ischemic stroke. Therefore, antioxidant supplementation is unlikely to be of clinical benefit to prevent ischemic stroke.

## Data Availability

The authors are grateful to the METASTROKE consortium and the FinnGen Biobank for making their summary statistics available for further studies. The analyses done in UK Biobank were done under project number 56340.

https://www.megastroke.org/

https://www.ukbiobank.ac.uk/

https://www.finngen.fi/en

