## Supplementary Table 1 for "Diet-derived antioxidants do not decrease the risk of ischemic stroke: a Mendelian Randomization Study in over 1 million participants"

Supplementary table 1: Ascorbate genetic instruments

| SNP | effect_allele | other_allele | eaf | beta | se | pval |
| --- | --- | --- | --- | --- | --- | --- |
| rs6693447 | T | G | 0.551 | 0.039 | 0.006 | 6.25E-10 |
| rs13028225 | T | C | 0.857 | 0.102 | 0.009 | 2.38E-30 |
| rs33972313 | C | T | 0.968 | 0.360 | 0.018 | 4.61E-90 |
| rs10051765 | C | T | 0.342 | 0.039 | 0.007 | 3.64E-09 |
| rs7740812 | G | A | 0.594 | 0.038 | 0.006 | 1.88E-10 |
| rs174547 | C | T | 0.328 | 0.036 | 0.007 | 3.84E-08 |
| rs117885456 | A | G | 0.087 | 0.078 | 0.012 | 1.70E-11 |
| rs2559850 | A | G | 0.598 | 0.058 | 0.006 | 6.30E-20 |
| rs10136000 | A | G | 0.283 | 0.040 | 0.007 | 1.33E-08 |
| rs56738967 | C | G | 0.321 | 0.041 | 0.007 | 7.62E-10 |
| rs9895661 | T | C | 0.817 | 0.063 | 0.008 | 1.05E-14 |
